# Primary care functional features in China and their relationship with health service outcomes: a 15-year mixed methods systematic review protocol

**DOI:** 10.1101/2025.04.11.25325626

**Authors:** Yang Wang, Hua Jin, Dehua Yu

**Affiliations:** Department of General Practice, Research Center for General Practice, Yangpu Hospital, School of Medicine, Tongji University, Shanghai, China; Shanghai General Practice and Community Health Development Research Center, Shanghai, China

## Abstract

**Introduction:** Globally, primary care (PC) features like accessibility, continuity, and coordination enhance health outcomes. In China, 940,000 public facilities delivered 4.266 billion PC visits in 2022 (50.68% of total visits), focusing on chronic disease management and public health. Unlike Western gatekeeper models, China’s PC lacks this structure, influencing its characteristics and effectiveness.

**Objective:** This review synthesises China’s PC functional features and their relationship with health service outcomes since 2009, combining quantitative and qualitative evidence.

**Methods:** Using the JBI mixed-methods framework, we will search PubMed, Embase, Web of Science, Google Scholar, CNKI, and Wanfang Data for studies (quantitative, qualitative, mixed-methods). Two reviewers will screen and assess quality with JBI tools (>50% threshold). A convergent segregated approach integrates quantitative data (feature strength, associations with outcomes) with qualitative insights (stakeholder views on features and effects).

**Discussion:** This review may shed light on the functional features of China’s primary care system and their relationship with health service outcomes. The synthesized evidence could inform clinical practice, health service delivery, and health policy regarding the health service process of primary care in China, while also identifying areas for future research.

## Background

Primary care, including family doctor services and those delivered by general practitioners, is known by various names but exhibits substantial overlap in clinical scope and modes of provision and access. Globally, leading theories on primary care quality assessment, rooted in North American and European research (e.g., Starfield’s framework), emphasize that evaluating functional features—such as accessibility, comprehensiveness, continuity, and coordination—is more critical than focusing solely on clinical technical quality^1,2^. Extensive evidence from these regions demonstrates a strong correlation between enhanced functional features and improved health service outcomes, such as better chronic disease control and patient satisfaction, making their assessment a core component of primary care evaluation^3-8^.

In China, primary care refers to essential medical services underpinning the healthcare system, as defined by legislation in 2019^9^. These services are delivered through approximately 940,000 publicly managed facilities, such as community health centers and township hospitals, centrally planned and funded by the government. With an aging population and rising chronic disease burden, these facilities provided 4.266 billion consultations in 2022 (50.68% of total healthcare visits, per the 2023 China Health Statistics Yearbook), highlighting their critical role in sustaining equitable access and population health^10^. Unlike the U.S., where international theories on functional features originated, China’s primary care focuses narrowly on common illnesses and chronic disease management (e.g., diabetes, hypertension), incorporates proactive community-based public health services (e.g., patient education, follow-ups), and operates without a gatekeeping system, competing with specialized hospital departments in a market-driven environment^9,11^. This unique blend of government oversight and market dynamics, shaped by China’s social and demographic context, may foster distinct functional features influencing health service outcomes.

Chinese researchers have adapted tools like the Primary Care Assessment Tool (PCAT)^12^, General Practice Assessment Questionnaire (GPAQ)^13^, and Person-Centered Primary Care Measure (PCPCM)^14^, validating them locally. However, quantitative data from these tools show inconsistent results, complicated by local practice variations^15^, while qualitative studies suggest context-specific features like consultation time and perceived respect^16-18^. Despite these efforts, no systematic review has integrated quantitative and qualitative evidence to clarify how these unique functional features relate to health service outcomes. Our preliminary search of PubMed, Google Scholar, CNKI, and PROSPERO found no existing or ongoing mixed-methods systematic reviews (MMSRs) on this topic.

This mixed-methods systematic review aims to address this gap by examining the functional features of China’s primary care and their associations with health service outcomes since 2009, following China’s healthcare reforms^19^. Quantitatively, it will assess the intensity levels of these features and their associations with outcomes. Qualitatively, it will explore stakeholders’ interpretations of these features and potential pathways of influence. By integrating these findings, it will provide evidence to guide practices, tools, and policies for measuring primary care quality in China and similar low- and middle-income countries.

### Review Question

This mixed-methods systematic review aims to summarize the functional features of China’s primary care and their relationship with health service outcomes, analyzing studies conducted in China’s primary care context from 2009 to March 31, 2025. It addresses four research questions: Qualitative: How do patients and service providers perceive the functional features of China’s primary care? What potential pathways might link these features to health service outcomes? Quantitative: What are the intensity levels of these functional features? How are they associated with health service outcomes?

Through separate syntheses of qualitative and quantitative evidence, followed by their integration, this review examines stakeholders’ experiences and perceptions of the performance of functional features, their reflections on the intensity of these features, and the potential relationships between these features and health service outcomes in primary care across mainland China.

### Inclusion and Exclusion Criteria

#### Inclusion Criteria

Studies must

1. Involve patients accessing primary care services in community settings or service providers (e.g., general practitioners, family doctor teams) in China’s primary care system.
2. Be published or completed between January 1, 2009, and March 31, 2025.
3. Be conducted in mainland China.
4. Be original research articles (e.g., journal articles, conference papers) or gray literature (e.g., theses).
5. Use quantitative designs (e.g., cohort, cross-sectional), qualitative designs (e.g., interviews, focus groups), or mixed-methods designs with clearly distinguishable qualitative and quantitative components.
6. Quantitative Studies: Assess intensity levels of primary care functional features (e.g., accessibility, continuity, coordination) as a whole or for specific features individually, and their associations or correlations with health service outcomes (e.g., population health, medical cost, patient satisfaction).
7. Qualitative Studies: Explore stakeholders’ perceptions of primary care functional features (e.g., accessibility, continuity) and the potential pathways that might link these features to health service outcomes.

#### Exclusion Criteria

Studies are excluded if they:

1. Do not explicitly or implicitly address primary care functional features (e.g., accessibility, continuity) or their sub-components.
2. Are non-empirical (e.g., reviews, editorials, protocols, case reports).
3. Were published or completed before January 1, 2009.
4. Were conducted outside mainland China.
5. Lack sufficient data or methodological detail for analysis (e.g., abstracts only).

## Method

This mixed-methods systematic review will follow the Joanna Briggs Institute (JBI) methodology for mixed-methods reviews, employing a convergent segregated approach to synthesize and integrate qualitative and quantitative evidence^20^. The process will include searching for relevant studies, selecting them based on predefined criteria, appraising their quality, extracting data, and synthesizing and integrating findings to address the review questions. The review will be registered with PROSPERO (No. CRD420251028411).

### Search Strategy

The search strategy will aim to identify published and unpublished studies addressing the review questions and inclusion criteria. It was developed iteratively: an initial search in PubMed and China National Knowledge Infrastructure (CNKI) identified articles on China’s primary care functional features, extracting text words (e.g., ‘primary care,’ ‘accessibility,’ ‘pathway’) and index terms (e.g., MeSH: ‘Primary Health Care’; CNKI: ‘基层医疗’ and ‘全科医疗’). These were combined with methodological terms (e.g., ‘quantitative,’ ‘qualitative’) and refined by a librarian to cover population (patients, providers), functional features (e.g., coordination), pathways, and study designs (quantitative, qualitative, mixed-methods).

Searches will be conducted across PubMed, Embase, Web of Science, CNKI, Wanfang Data, and Google Scholar (first 50 pages, justified by diminishing relevance), targeting English and Chinese studies from January 1, 2009, to March 31, 2025. Original research and gray literature (e.g., theses from CNKI and Wanfang, supplemented by hand-searching policy reports) will be included. Detailed strategies, tailored to each database’s syntax and excluding non-empirical types (e.g., reviews), are in Appendix 1. Reference lists of appraised studies will be screened for additional sources.

### Study Selection

After executing the search strategies (Appendix 1), citations will be uploaded to Rayyan (Rayyan Systems Inc.)^21^ for deduplication and screening. Two independent reviewers, Yang Wang and Hua Jin, will screen titles and abstracts against the inclusion criteria, retrieving potentially relevant full texts for eligibility assessment using the inclusion and exclusion criteria. Exclusions will be documented with reasons and reported in the final review. Disagreements at either stage (title/abstract or full-text) will be resolved through discussion or by a third reviewer (Dehua Yu). The search and selection process will be detailed in the final review, presented via a PRISMA flow diagram^22^.

### Assessment of Methodological Quality

Included studies will be appraised for methodological quality using JBI’s critical appraisal tools to study design: quantitative studies (e.g., cohort, cross-sectional) with checklists for analytical cross sectional studies^23^ or cohort studies^24^, qualitative studies with checklist for qualitative research^25^, and mixed-methods studies with both tools applied separately to distinct components. Each checklist item will be scored (0 = not met, 0.5 = partially met, 1 = fully met), with total scores as percentages: low (<50%), moderate (50%-75%), or high (>75%). Only studies scoring >50% will be synthesized, prioritizing credible evidence; lower-scoring studies may inform background context. Two reviewers, Yang Wang and Hua Jin, will independently assess quality, resolving disagreements via discussion or a third reviewer (Dehua Yu). Authors will be contacted for missing data (noted as unavailable if no response within 2 weeks). Results will be reported narratively and tabulated in the final review.

### Data Extraction

Data will be extracted from included studies by two independent reviewers, Yang Wang and Hua Jin, using a pre-designed Excel table (Appendix 2) tailored to the review questions. Given that functional features may serve as exposures or outcomes across studies, we adapted the Donabedian Structure-Process-Outcome (SPO)^26^ model instead of traditional PICO elements. In this adaptation, “Structure” is defined as factors that may positively or negatively influence functional features (corresponding to the “S” in SPO), “Functional Features” as a set of service characteristics arising from the primary care process that affect process quality (corresponding to the “P” in SPO), and “Outcomes” as the resulting health service effects (corresponding to the “O” in SPO). For quantitative studies, data will include study characteristics (e.g., author, year), population, design, structural factors (e.g., family doctor contract services), functional feature intensity levels (e.g., overall PCAT scores or accessibility on a 5-point Likert scale, with maximum values), comparison conditions (e.g., high vs. low intensity), health service outcomes (e.g., satisfaction), and measurement tools. For qualitative studies, data will cover study characteristics, population, context (e.g., community health centers), stakeholders’ perceptions of their experiences with functional features (e.g., coordination), and potential pathways that might link these features to health service outcomes (e.g., trust mechanisms). Mixed-methods studies will have both data types extracted separately where distinguishable, supporting the convergent segregated approach. Disagreements will be resolved via discussion or a third reviewer (Dehua Yu). Authors will be contacted for missing data (coded ‘N/A’ if unavailable after 2 weeks). Data will be finalized in Excel for synthesis.

### Data Synthesis and Integration

This review will use a convergent segregated approach, per JBI mixed-methods methodology, to synthesize and integrate data addressing China’s primary care functional features and their relationship with health service outcomes. Data extracted via Appendix 2 will be synthesized separately for quantitative and qualitative evidence, then integrated, with findings presented in tables and narrative form.

Quantitative synthesis will address:

#### Intensity Levels of Functional Features

Where feasible, meta-analysis will pool intensity scores separately for each measurement tool (e.g., overall PCAT or PCPCM scores) using Stata, with heterogeneity assessed via chi-squared and I² statistics (random-effects model if I² > 50%). If significant population differences (e.g., rural vs. urban patients, patients vs. providers) contribute to heterogeneity, subgroup analyses by population type will be conducted where data permit. Sub-dimension scores (e.g., accessibility, continuity) will be synthesized separately within each tool where data allow, using similar methods. If meta-analysis is precluded by heterogeneity (e.g., diverse scales, population variability), descriptive statistics (e.g., medians, ranges by tool, sub-dimension, and population) or narrative synthesis will summarize intensity levels.

#### Associations with Health Service Outcomes

Meta-analysis will estimate effect sizes (e.g., odds ratios from controlled analyses) where data permit, pooling studies with consistent exposure and outcome tools (e.g., PCAT to Likert satisfaction). Subgroup analyses by tool will be conducted if sufficient studies exist for diverse tools (e.g., PCAT vs. PCPCM); additional subgroup analyses by population type (e.g., patients vs. providers, rural vs. urban) will be performed if heterogeneity suggests population-driven differences. If pooling is inappropriate due to heterogeneity (e.g., tool differences, population variability), descriptive statistics (e.g., OR ranges by tool and population) or narrative synthesis will describe associations.

Qualitative synthesis will examine:

#### Stakeholders’ Perceptions of Experiences with Functional Features

Qualitative data on stakeholders’ perceptions (e.g., patients’ and providers’ experiences with accessibility or continuity) will be synthesized using meta-aggregation^25^, a JBI method that extracts findings from studies, groups them by similarity in meaning, and produces synthesized statements to represent shared experiences. This process, supported by MAXQDA software for coding and data management, will be applied where sufficient data are available. If meta-aggregation is not feasible (e.g., due to limited studies or diverse foci), thematic analysis will identify recurring themes, or narrative synthesis will describe perceptions descriptively. Differences in stakeholder perspectives (e.g., rural vs. urban patients, community vs. hospital-based providers) will be explored to capture diverse experiences.

#### Potential Pathways to Health Service Outcomes

Qualitative evidence on pathways linking functional features to outcomes (e.g., trust mechanisms or care coordination effects) will be synthesized using meta-aggregation where data permit, categorizing findings into synthesized statements with MAXQDA’s assistance. If meta-aggregation is unsuitable (e.g., due to sparse or heterogeneous data), thematic analysis will group pathways into themes, or narrative synthesis will provide a descriptive summary. Two reviewers (Yang Wang and Hua Jin) will independently conduct the synthesis, resolving disagreements via discussion or with a third reviewer (Dehua Yu). Results will be tabulated (e.g., by pathway category) and narrated, informing the integration phase by elucidating mechanisms behind quantitative associations.

### Integration of Quantitative and Qualitative Findings

A juxtaposition table (Appendix 3) will integrate quantitative and qualitative syntheses, focusing on functional features and their influence on health service outcomes. It will assess consistency, contradictions, or complementarity (e.g., how intensity aligns with perceptions, or pathways explain associations), avoiding pre-specified examples to maintain neutrality. Integrated results will address the review questions, presented in tables and narrative form.

### Supplementary Analysis of Measurement Tools

Characteristics of measurement tools (e.g., PCAT dimensions, Likert scale properties) will be summarized descriptively or narratively as supplementary context, reported in tables and text, but excluded from the primary integration.

## Data Availability

All data produced in the present work are contained in the manuscript

## Acknowledgements

This review is conducted as part of Yang Wang, Hua Jin and Dehua Yu’s research activities, with no external funding or contributions to acknowledge.

## Author Contributions

Conceptualization, Y.W.; Methodology, Y.W; Data curation, Y.W and H.J; Formal analysis, Y.W.; Funding acquisition, Y.W and H.J; Project administration, Y.W and D.Y; Resources, Y.W and D.Y; Supervision, D.Y; Validation, Y.W; Writing—original draft, Y.W; Writing—review and editing, Y.W, H.J and D.Y. All authors have read and agreed to the published version of the manuscript.

## Ethics and Dissemination

As a systematic review of published data, this study requires no ethical approval. Findings will be disseminated through peer-reviewed publication and conference presentations to inform primary care research and policy in China.

## Amendments

Any amendments to this protocol will be documented with rationale and updated in PROSPERO as needed.

## Appendix 1 Search Strategies

These search strategies were developed to identify relevant studies for this mixed-methods systematic review on China’s primary care functional features and their relationship with health service outcomes. Searches were conducted across six databases: PubMed, Embase, Web of Science (WOS), Google Scholar, China National Knowledge Infrastructure (CNKI), and Wanfang Data. Each strategy was tailored to the database’s syntax and capabilities, targeting studies published or completed between January 1, 2009, and March 31, 2025. Results are reported as of the search execution date, to be updated upon completion (e.g., March 30, 2025).

### PubMed

#### Search String

((“Primary Health Care”[MeSH Terms] OR “General Practice”[MeSH Terms] OR “primary care”[tiab] OR “primary healthcare”[tiab] OR “basic medical”[tiab] OR “general practice”[tiab] OR “community health”[tiab]) AND (“functional features”[tiab] OR “functional characteristic*”[tiab] OR “functional attribute*”[tiab] OR “function feature*”[tiab] OR “function attribute*”[tiab] OR “functional trait*”[tiab] OR “functional propert*”[tiab] OR “service feature*”[tiab] OR “service characteristic*”[tiab] OR “service attribute*”[tiab] OR “quality”[tiab] OR “process quality”[tiab] OR “process characteristic*”[tiab] OR “performance”[tiab] OR “first contact”[tiab] OR “accessibility”[tiab] OR “comprehensiveness”[tiab] OR “continuity”[tiab] OR “coordination”[tiab]) AND (“China”[MeSH Terms] OR “China”[tiab] OR “Chinese”[tiab]) AND (“quantitative”[tiab] OR “qualitative”[tiab] OR “mixed methods”[tiab] OR “survey”[tiab] OR “interview”[tiab] OR “cohort”[tiab] OR “cross-sectional”[tiab] OR “focus group”[tiab] OR “statistical”[tiab]) AND (“methods”[tiab] OR “methodology”[tiab]) NOT (“Review”[Publication Type] OR “Comment”[Publication Type] OR “Editorial”[Publication Type] OR “Practice Guideline”[Publication Type] OR “Case Reports”[Publication Type] OR “Clinical Trial Protocol”[Publication Type] OR “Letter”[Publication Type])) AND (“2009/01/01”[PDAT] : “2025/03/31”[PDAT]) AND English[lang]

Notes: Language: English; Time Range: January 1, 2009, to March 31, 2025

### Embase

#### Search String

(‘primary health care’:ti,ab,kw OR ‘general practice’:ti,ab,kw OR ‘primary care’:ti,ab,kw OR ‘primary healthcare’:ti,ab,kw OR ‘basic medical’:ti,ab,kw OR ‘general practice’:ti,ab,kw OR ‘community health’:ti,ab,kw OR ‘primary health care’/exp OR ‘general practice’/exp) AND (‘functional feature*’:ti,ab,kw OR ‘functional characteristic*’:ti,ab,kw OR ‘functional attribute*’:ti,ab,kw OR ‘function feature*’:ti,ab,kw OR ‘function attribute*’:ti,ab,kw OR ‘functional trait*’:ti,ab,kw OR ‘functional propert*’:ti,ab,kw OR ‘service feature*’:ti,ab,kw OR ‘service characteristic*’:ti,ab,kw OR ‘service attribute*’:ti,ab,kw OR ‘service quality’:ti,ab,kw OR ‘process quality’:ti,ab,kw OR ‘process characteristic*’:ti,ab,kw OR ‘performance’:ti,ab,kw OR ‘first contact’:ti,ab,kw OR ‘accessibility’:ti,ab,kw OR ‘comprehensiveness’:ti,ab,kw OR ‘continuity’:ti,ab,kw OR ‘coordination’:ti,ab,kw) AND (‘China’:ti,ab,kw OR ‘Chinese’:ti,ab,kw OR ‘China’/exp) AND (‘quantitative’:ti,ab,kw OR ‘qualitative’:ti,ab,kw OR ‘mixed methods’:ti,ab,kw OR ‘survey’:ti,ab,kw OR ‘interview’:ti,ab,kw OR ‘cohort’:ti,ab,kw OR ‘cross-sectional’:ti,ab,kw OR ‘focus group’:ti,ab,kw OR ‘statistical’:ti,ab,kw) NOT (‘review’:pt OR ‘comment’:pt OR ‘editorial’:pt OR ‘practice guideline’:pt OR ‘case report’:pt OR ‘clinical trial protocol’:pt OR ‘letter’:pt)

Notes: Language: English; Time Range: January 1, 2009, to March 31, 2025

### Web of Science (WOS)

#### Search String

TS=(“Primary Health Care” OR “General Practice” OR “primary care” OR “primary healthcare” OR “general practice” OR “community health”) AND TS=(“functional feature*” OR “functional characteristic*” OR “functional attribute*” OR “function feature*” OR “function attribute*” OR “functional trait*” OR “functional propert*” OR “service feature*” OR “service characteristic*” OR “service attribute*” OR “service quality” OR “process quality” OR “process characteristic*” OR “performance” OR “first contact” OR “accessibility” OR “comprehensiveness” OR “continuity” OR “coordination”) AND TS=(“China” OR “Chinese”) AND TS=(“quantitative” OR “qualitative” OR “mixed methods” OR “survey” OR “interview” OR “cohort” OR “cross-sectional” OR “focus group” OR “statistical”) AND AB=(“methods”) NOT TS=(“review” OR “comment” OR “editorial” OR “guideline” OR “case report” OR “protocol” OR “letter”)

Notes: Language: English; Time Range: January 1, 2009, to March 31, 2025

### Google Scholar (limited to first 50 pages)

#### Search String

“primary care” OR “primary healthcare” OR “general practice” OR “community health” “functional feature*” OR “functional characteristic*” OR “functional attribute*” OR “service feature*” OR “service characteristic*” OR “first contact” OR “accessibility” OR “comprehensiveness” OR “continuity” OR “coordination” “China” OR “Chinese” “study” -”review” -”comment” -”editorial” -”guideline” -”case report” -”protocol” -”letter”

Notes: Language: English; Time Range: January 1, 2009, to March 31, 2025; Results: Limited to first 50 pages (approximately 500 results, justified by relevance decline after initial pages)

Note: Google Scholar’s result count varies; focus is on the first 50 pages due to practical constraints.

### China National Knowledge Infrastructure (CNKI)

#### Search String

TKA=(“全科医学” OR “基层医疗” OR “社区卫生” OR “初级保健” OR “初级卫生保健” OR “基本医疗” OR “ 家庭医生”) AND TKA=(“质量” OR “绩效” OR “功能” OR “体验” OR “浈量” OR “评估” OR “服务” OR “特性” OR “属性” OR “首诊”) AND AB=“方法” NOT TKA=(“综述” OR “评论” OR “述评” OR “指南” OR “病例报告” OR “方案”)

Notes: Language: Chinese; Time Range: January 1, 2009, to March 31, 2025

### Wanfang Data (451 results)

#### Search String

主题=(“全科医学” OR “基层医疗” OR “社区卫生” OR “初级保健” OR “初级卫生保健” OR “基本医疗” OR “ 家庭医生”) AND 主题=(“功能特征” OR “服务特征” OR “征程质量” OR “服务质量” OR “首诊” OR “可及性” OR “全面性” OR “连续性” OR “协调性”) AND 主题=(“中国”) AND 摘要=“方法” NOT 主题=(“综述” OR “评论” OR “述评” OR “指南” OR “病例报告” OR “方案”)

Notes: Language: Chinese; Time Range: January 1, 2009, to March 31, 2025

## Appendix 2 Draft Data Extraction Table

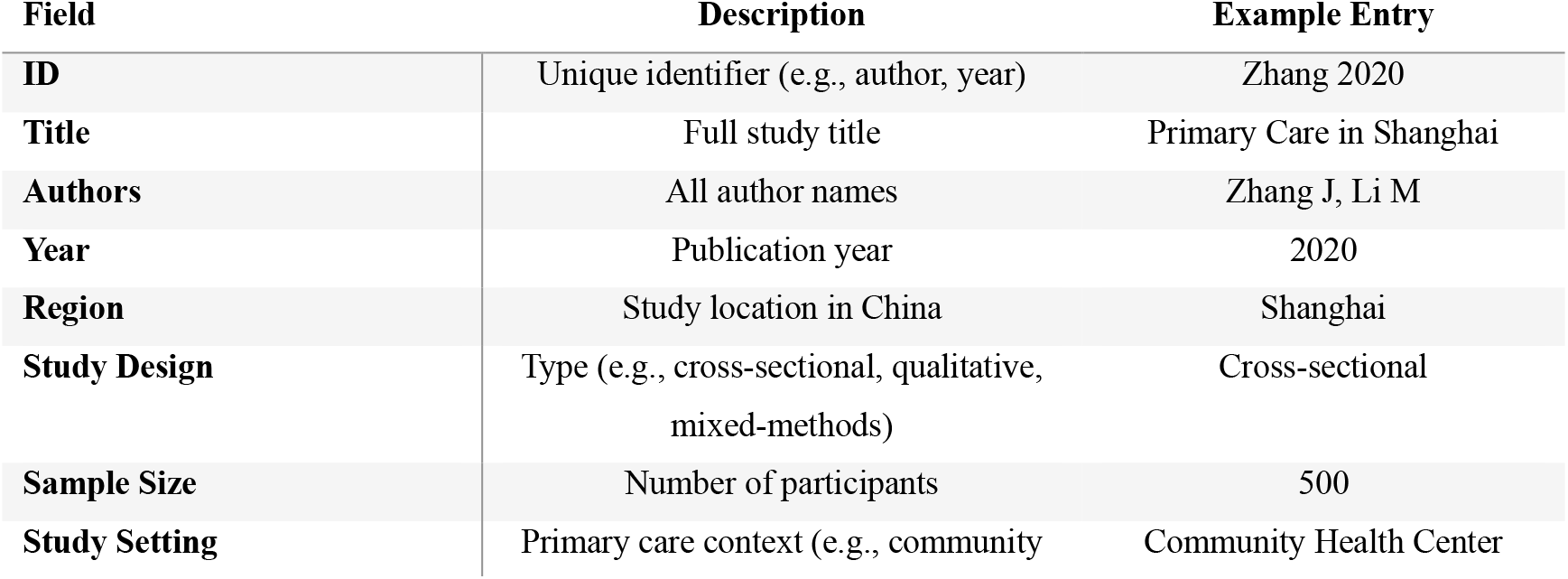
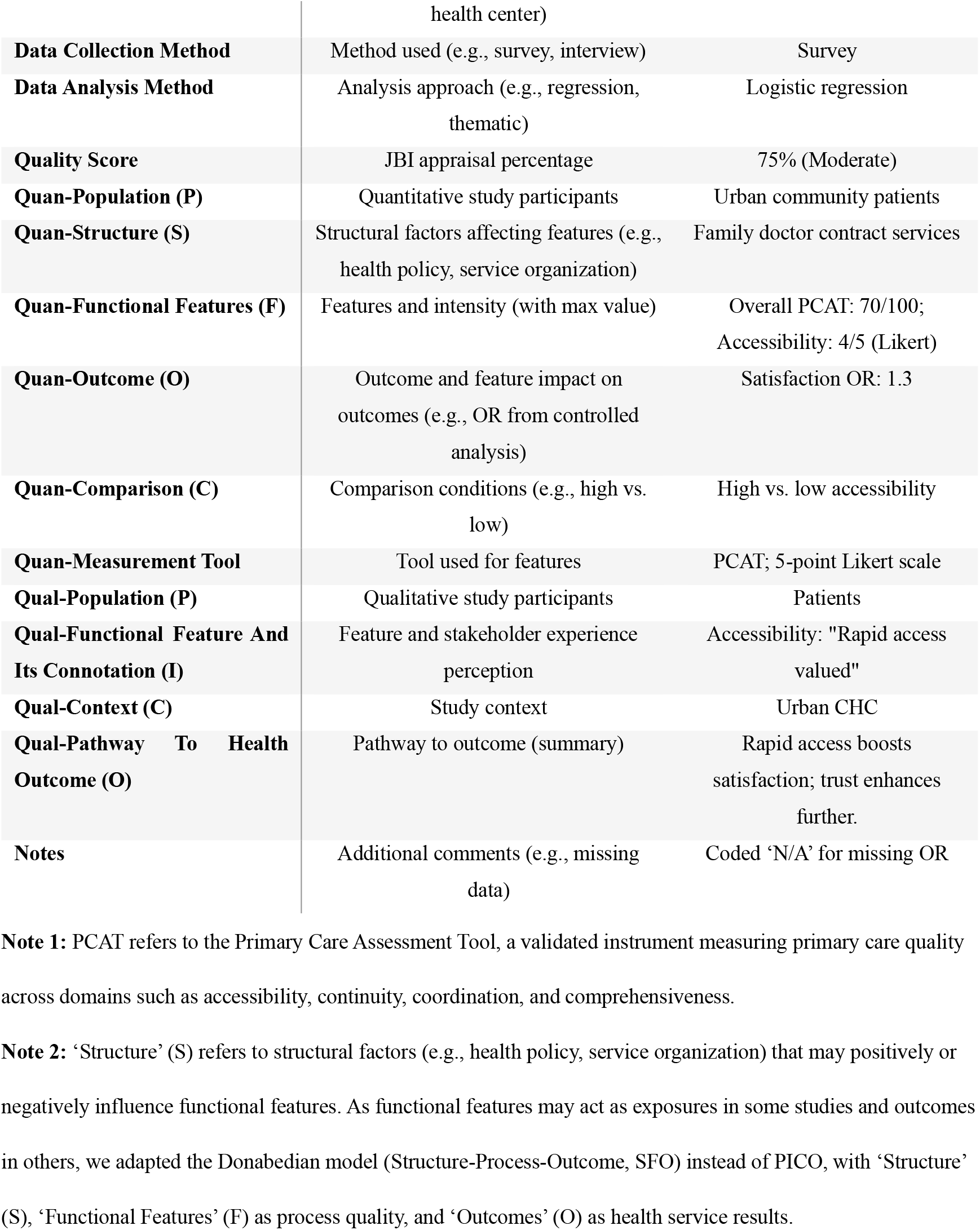

## Appendix 3 Sample Juxtaposition Table for Integration

Appendix 3: Sample Juxtaposition Table for Integration
| Category | Feature | Quantitative Synthesis | Qualitative Synthesis | Integration Findings |
| --- | --- | --- | --- | --- |
| **Functional Features (F)** | Accessibility | Intensity TBD (e.g., Likert scores) | Perception TBD (e.g., access experience) | TBD (e.g., consistency expected) |
| **Functional Features (F)** | Continuity | Intensity TBD (e.g., PCAT scores) | Perception TBD (e.g., trust experience) | TBD (e.g., consistency expected) |
| **Functional Features (F)** | Overall | Intensity TBD (e.g., overall PCAT) | Perception TBD (e.g., overall experience) | TBD (e.g., consistency expected) |
| **Influence on Outcomes (O)** | Accessibility | Impact TBD (e.g., association with outcome) | Pathway TBD (e.g., access mechanism) | TBD (e.g., complementarity expected) |
| **Influence on Outcomes (O)** | Continuity | Impact TBD (e.g., association with outcome) | Pathway TBD (e.g., trust mechanism) | TBD (e.g., complementarity expected) |
| **Influence on Outcomes (O)** | Overall | Impact TBD (e.g., combined association) | Pathway TBD (e.g., overall mechanism) | TBD (e.g., complementarity expected) |

**Note:** TBD = To Be Determined; specific results will be determined post-synthesis.

